# Barriers to climate change and health research in India: A qualitative study

**DOI:** 10.1101/2023.01.26.23284955

**Authors:** Shreya S Shrikhande, Sonja Merten, Olga Cambaco, Tristan Lee, Ravivarman Lakshmanasamy, Martin Röösli, Mohammad Aqiel Dalvie, Jürg Utzinger, Guéladio Cissé

## Abstract

Almost a quarter of the global burden of disease and mortalities is attributable to environmental causes, the magnitude of which is projected to increase in the near future. Evidence informed policies and interventions are a key element in the adaptation response for countries. However, in many low- and middle-income settings, there remains a large gap in the synthesis of evidence on climate-sensitive health outcomes. In India, now the world’s most populous country, little remains known about the impacts of climate change on various health outcomes. In light of India’s vulnerability to climate change, the growing population and the high burden of disease, it is imperative for public health professionals to engage in the climate action, and to understand the challenges they face, particularly with regard to barriers in conducting environmental health research. In this study, we employed key informant interviews to understand the perceived research barriers amongst health professionals, including medical researchers, and professionals involved in environmental policies and planning in Puducherry, India. The findings were analysed using data driven qualitative thematic analysis to elaborate the major perceived barriers to conducting environmental health research. Challenges in data collection systems and accessing data was the major barrier along with the need for strengthening technical and methodological research capacity. Participants described working in the backdrop of insufficient prioritization and knowledge on the wide range of impacts of climate change on health, both in the policy context and amongst scientists, which was also perceived to be a challenge in conducting environmental health research. Finally, limited resources to conduct research and the tendency to focus on conventional climate related health outcomes were also seen as challenges to advancing research on this topic. In the context of the paucity of data on environmental health from India, despite recognised climate change related health vulnerabilities, these findings could contribute to the development and improvement of relevant interventions conducive to a strong research environment.

**Key message:** *What is already known on this topic?:* Climate change has been linked to a range of adverse health outcomes globally. There is a growing body of research studying the associations between climate variables and various health outcomes. However, it remains poorly studied in India, which has a high vulnerability to climate impacts. It is important to understand what public health professionals perceive as barriers on the matter and their needs in order to better elucidate the health risks of climate change and improve the public health response to adapt to them.

*What this study adds?:* We identify three key barriers faced by public health professionals as key stakeholders, namely: (i) political and institutional barriers;(ii) education and awareness barriers; and (iii) technical research barriers. We show there is a need, from the professionals perspective, to improve community and political awareness on climate change and health; improve technical research capacity and collaboration amongst researchers; and improve health surveillance and access to health data for research.

*How this study might affect research, practice or policy:* This study identifies crucial challenges faced in conducting environmental health research by public health professionals. Therefore, the findings can be used to further elaborate and address these challenges, in order to further motivate the professionals, strengthen the environmental health research capacity and improve understanding of health vulnerabilities and risks attributed to climate change in India.

## INTRODUCTION

An ever-growing body of research has irrefutably shown the global health impacts of climate change through both direct and indirect exposure pathways [1, 2]. Multiple risk and vulnerability factors determine the population resilience and adaptive capacity, from socio-political, demographic and biological factors to infrastructure, urban planning, health information systems and health workforce [2, 3]. Given the regional variations in climate systems, the health impacts of climate change differ between and within countries and communities, mediated by interconnected socio-economic and environmental determinants of health [4, 5]. Non-communicable diseases (NCDs), such as respiratory diseases, cardiovascular diseases (CVDs), mental health conditions, have been recognized as growing climate-sensitive health outcomes, in addition to other communicable diseases like vector- and water-borne diseases and malnutrition [3, 6].

With the rapid pace of climate change, the health impacts attributable to it are also projected to increase [7]. Strengthening the adaptive capacity of countries is therefore an essential component of the climate change response [8]. Timely public health interventions can do much to protect population health from the potential adverse health impacts of climate change [9]. Low- and middle-income countries (LMICs), such as India, remain disproportionately affected by climate impacts, with a critical need to strengthen the healthcare response to climate impacts [10, 11]. One of the key steps in the regional or local adaptation response is assessing the true burden of the health impacts within the population of that location [12]. However, owing to the complexity of the relationship between climate change and health, identifying and estimating this association remains one of the biggest global and environmental health challenges, especially in LMICs [11].

In India, the existing health and social disparities within the population make it one of the most vulnerable to climate change impacts, compounded by climatic diversity [13-16]. There have been recent efforts from the Government of India to focus on climate change and health, as evinced by the recent addition of a health mission to the National Action Plan on Climate Change (NAPCC). This led to the formulation of the National Action Plan on Climate Change and Human Health (NAPCCHH) and the drive for State Action Plans for Climate Change and Human Health (SAPCCHH) [17, 18]. While the government recognizes in those official document several diseases as climate-sensitive, public health engagement, action and research on health impacts of climate change have been curiously limited in India, especially in light of the magnitude of climate impacts to which it is vulnerable [19, 20].

Medical and public health professionals play an important role in researching, managing and responding to climate change impacts on health. Along with being considered credible sources of information, these groups of professionals also have the capacity for scientific inquiries into the climate change attributable impacts of health [21-24]. Globally, there is an acknowledged need to train the health workforce to engage on, study and manage health impacts of climate change. As there are currently gaps in this domain, it is therefore particularly important to understand how this group perceive the needs and the barriers for their appropriate level of engagement and action any perceived research needs and barriers identified by this group [25, 26].

The aim of this study is to understand some of the contextual barriers to environmental health action-research amongst two relevant professional groups in Puducherry, India. We focused our study on: (i) medical professionals, both in active research and practicing; and (ii) members of the Department of Science, Technology and Environment working on climate change in Puducherry. As this study is a part of a larger project on CVDs and climate change in India, we also highlighted the CVD specific challenges and barriers to conducting research. As the focus of our paper is to understand challenges, research enablers have not been highlighted.

## METHODS

### Study setting

This study employed key informant interviews following a semi-structured interview guide. The methods have been described in detail elsewhere [27]. Briefly, the focus of our study was Puducherry district, which lies on the south-eastern coast of India, with a population of 950,289, as per the Government of India 2011 Census [28]. Puducherry has one main State government run tertiary care hospital and medical college, several private clinics and primary care health centres. It is also home to the Central Government Jawaharlal Institute of Postgraduate Medical Education and Research (JIPMER), an ‘Institute of National Importance’ and tertiary care referral hospital. Within the Department of Science, Technology and Environment (DSTE), there also exists a specialized Puducherry Climate Change Cell with the aim to integrate knowledge about climate change and facilitate the NAPCC implementation, including the state specific Action Plan [29].

### Data collection and analysis

A total of sixteen semi-structured interviews were conducted between January 2022 and March 2022 in Puducherry and virtually over Zoom for two participants. Using purposive sampling based on prior connections followed by snowball sampling, we invited medical professionals (research or practicing) and DSTE staff working on the Puducherry State Action Plan for Climate Change (hereon referred to as environmentalists). Interviews continued until information saturation was reached in the interviews or we had interviewed all the relevant target participants, as in the case of the DSTE staff. Practicalities such as participant schedules and ongoing COVID-19 restrictions also influenced our informant recruitment.

There was an equal balance between research-engaged professionals from the medical and environmental field combined and practicing doctors. Eleven of the participants had a medical background and were actively engaged as practicing doctors or researchers. Within the doctors, we mainly targeted cardiologists, emergency medicine or general medicine physicians who were involved in areas relevant to our study. The majority of the participants was male, with only three females, out of which only one had a medical background. The participant profile is presented in the Supplementary material Table S3 and further described in [27].

The interviews lasted between 15 to 50 minutes and were audio recorded with informed consent using a simple voice recorder and field notes were taken to optimize the interview guide and note key topics to minimize bias. We used an a priori developed interview guide with broad and open-ended questions to allow participants to freely bring up and discuss relevant topics. All interview recordings were assigned a number prior to transcription to ensure anonymity throughout the analysis process. Verbatim transcription and analysis was done using the MaxQDA software by S.S.

For the analysis, we followed a combination of deductive and inductive thematic analysis was used as described by Gale et al [30]. Broad themes were developed based on the aim, framework and interview guide. During analysis, major themes were inductively developed for emerging topics, which we then clearly defined. After familiarization with the transcripts, an initial codebook was developed from coding the three interviews with the richest data; the remaining interviews were indexed and coded further. The codes were classified into categories, sub-themes and themes. The final analytical matrix included three themes. S.S. and T.L. independently validated the codebook and agreed on the final framework matrix that considered all relevant codes. The matrix was then used to chart relevant quotes supporting our findings and draw comparisons between participants.

The conceptual framework for climate change risk perceptions developed by van Eck et al [31] and the framework for health inequalities proposed by Rudolph et al [32] were used as a base for our analytical framework, shown in Figure 1. While there are three major themes, this paper focuses only on the theme of ‘Institutional determinants’. The findings from the two other themes have been elaborated elsewhere. Within the context of this paper, ‘institution’ is used as a broad term covering all governmental structures including policy, education, and occupation. We identify how these determinants can be perceived as barriers to environmental health research.

**Figure 1:**
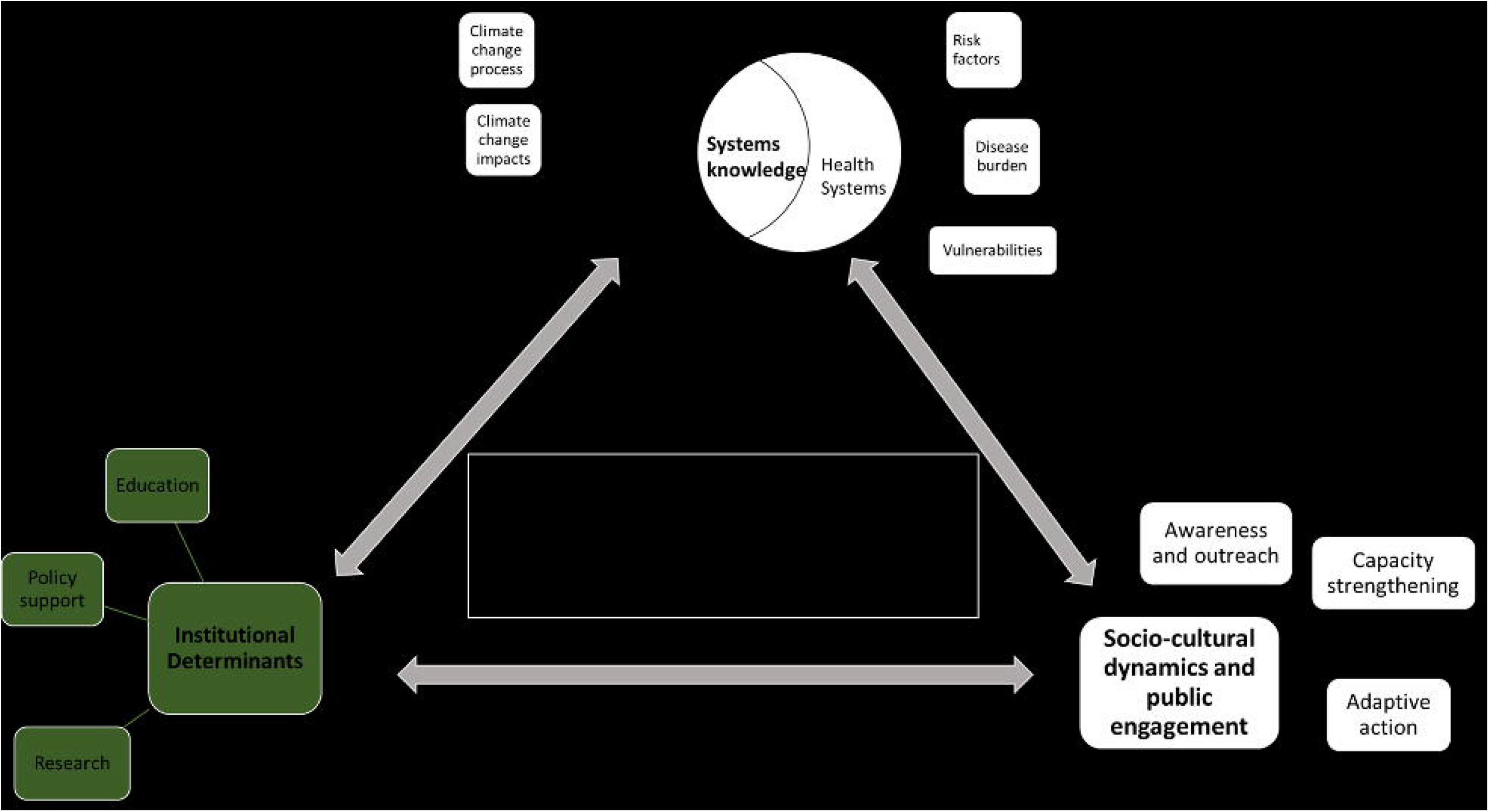
A framework for health adaptation action in the context of climate change based on level of knowledge, perceived health risks, policy and institutional support and public engagement. The circled part highlights the thematic areas we focus on in this work, namely institutional determinants and its challenges.

### Ethical consideration

There was no prior relationship between the researcher and participant. Before the interview, the researcher went over the informed consent form, which was then signed by both parties. The participants were aware of their right to withdraw from the study at any time, and they were provided the contact details of all the researchers involved in the project. All interviews were carried out by S.S with R.L being present as a passive observer for three interviews. Additionally, all the quotes presented in this analysis have been assigned only by serial number to ensure anonymity.

This study was approved by the Institute Ethics Committee (Human Studies) of the Indira Gandhi Medical College and Research Institute (A Govt. of Puducherry Institution); No. 318/IEC-31/IGM&RI/PP/2021 and by the Ethics Committee Northwest and Central Switzerland (EKNZ); Statement ID-AO_2020_00034. The methodology used in this project abided by the principles laid out in the Declaration of Helsinki and the COREQ checklist.

### Patient and public involvement

As we employed a combination of purposive and snowball sampling, some participants were involved in helping us identify suitable participants to interview. Beyond that, no members of the public were involved in the design, conduct, reporting or dissemination plan of our research.

## RESULTS

We first lay out the knowledge participants had regarding the institutional framework, including policies, for climate change and health followed by perceived institutional barriers to research, namely political, educational and technical barriers. As this study is part of a larger study examining climate change impacts on CVDs, we also chose to highlight CVD specific barriers.

### Institutional determinants: knowledge and perceptions

#### Limited knowledge and awareness on climate change and health related policies

We found limited awareness among the participants about climate change and health related policies, such as the NAPCC, NAPCCHH and SAPCCHH. Aside from the environmentalists, who worked on it, only three medical professionals who worked on one of the Action Plans were aware of it. Four participants expressed belief about the non-inclusion of climate change in disease specific policies and the lack of integrated climate change and health policies and guidelines.

*“Our country has different policy, environmental policy, health policy. But I have doubt whether health policy has any component of climate change. So, it needs to be incorporated in a health policy of national importance as well as the state, but currently, this element is not in place, that is my feeling.”* #8, Environmentalist

One of the environmentalist also mentioned challenges faced by the government in integrating climate change in development plans. The emphasis on socio-economic development was made apparent, which was countered by ongoing efforts to include climate change adaptation as co-benefits of development.

*“The challenge is that the government sectoral officers are not aware of how the climate adaptations need to be integrated into their developmental plans. Because they, whenever they plan for a project, they plan it from the socio economic development perspective.”* #7, Environmentalist

Health system vulnerabilities due to climate change in Puducherry were considered a growing challenge and highlighted by multiple doctors. Most participants expressed limited knowledge about climate change and health and agreed on the importance of education as the first step to increase awareness and research capacity. Several participants, especially in those working in hospitals, displayed support to reform existing guidelines to reflect health risks from climate change, conditional to more contextual research to inform these changes.

*“This climate and health is a very good topic. To be frank, we don’t have much knowledge about climate and health. … After knowing all these things, after getting an idea what to happen, we should have a plan, national policy or something, we should make a national policy so that the government can do whatever needed during the season. So I think this will be the first step.”* #13, Practicing physician

In addition, we found varying knowledge among participants about the health impacts of climate change. Vector borne diseases were most commonly associated with climate-sensitivity while other diseases like NCDs or CVDs were perceived to be primarily lifestyle related. When it came to research, very few participants mentioned having been part of environmental health studies or knowing about current research. The need for regional research for knowledge synthesis on health risks from climate change was expressed throughout the interviews.

### Political and institutional barriers

#### Disengaged leadership and low political prioritization of climate change and health

Political leadership that did not consider health impacts of climate change as a pressing matter was perceived as one of the barriers to conducting research on the topic. Rather than viewed as a cause for immediate concern, several participants mentioned how climate change was rather seen as future concern by policy makers as well as the general public. Given the perceived little awareness on health impacts, a few participants also mentioned the slim likelihood of decision makers actually being aware of it. One participant described the issue as being *“not mainstream enough”* to warrant focused work, contributing to the perceived low priority assigned to environmental health research.

Many participants felt that the governmental focus was inclined towards non-health impacts of climate change. The most pressing climate change impacts, which also influence research focus, were thought to be pollution, coastal sensitivity and natural resource depletion and degradation, especially in the context of Puducherry as a coastal region. Additionally, existing sectoral programs already running were seen as a hindrance to focusing on climate change related programs by one participant.

*“The problem is everybody has to understand at the level of the minister or the secretaries. They have to come forward…because so many programs are there. Not only about climate change, other programs are there so they do not focus much on (climate change) programs… Actually, what I have seen for the past 2-3 years, they don’t care much about climate.”#1, Practicing physician/policy advisor*

The challenges India faces from other vulnerabilities were perceived to outrank climate risks to health, including unmet nutritional and economic needs. Despite climate change being recognized as a health risk factor, there was a clear disconnect between on-paper government plans and practice when it came to environmental health research.

*“I’m an adviser to government of India on health related research. But we did discuss a lot of things but we did also touch upon climate-effects of climate on health…That was considered as an important topic, but we didn’t dwell much upon how to take it forward because there are more pressing problems.”* #15, Practicing physician/academic

#### Weak inter-departmental integration and co-ordination for climate change and health

The compartmentalization of topics within institutes or sectors was seen as a barrier to conducting inter-disciplinary research by the participants. One participant discussed the newly formed Puducherry Climate Change and Health Action Plan (2022), with the aim to bring together a multi-sectoral team under the leadership of the health ministry, to focus on health impacts of climate change.

However, apart from one participant, most others voiced a perceived need for an independent, coordinating body focused on environmental health, incorporating a research agenda. Partially, this was due to climate change being thought of an added responsibility for health professionals and vice versa for environmentalists, especially for those working in the government. As highlighted by a few participants, officials were likely to prioritize their primary work profile over the added responsibility of climate change and health research.

*“Especially government departments, they are loaded with a lot of work. Today, an officer comes in, he has to do his own work, not the work that other departments asks us to do…”* #9, Environmentalist

Several participants mentioned the Puducherry Climate Change cell created in response to address climate change impacts. However, despite that, one medical researcher mentioned the current difficulties in identifying the focal point of contact for climate change and health. Another concern in the existing scenario was inter-sectoral, collaborative research being dependant on higher officials being receptive to their employees researching a topic not entirely within the scope of their respective department. This was also viewed from the sense of improving coordination between the sectors, with a dedicated head of climate change and health, supported by relevant sectorial bodies.

*“Intersectoral body and there should be one decision maker. So now, everybody is like they are the leader in the particular sector, but if they need the support from other one, that coordination may be lacking…There won’t be any one dedicated person for the climate change. So they will be in charge of multiple departments. For example, somebody’s going to be in charge of immunization or the child health. So the priority will be child health obviously.”* #3, Medical doctor/academic

### Educational and informational barriers

#### Gaps in climate change and health in higher education curricula

One of the strongest emergent themes was the need for environmental health education, either by incorporating climate change in the health curriculum or health impacts of climate change in the environmental curriculum in universities and schools. The prevalent feeling was the source of climate change and health literacy needs to be from multiple sources, with formal education being the most important one. Most participants also felt that at present there was a disconnect between environmental and health education, as a result of which there was a relatively low level of awareness on climate change impacts on health.

*“Education system need to be addressed from beginning…Even the medical college students who are completing five years courses, I do not see any syllabus which contains impact on health by the climate change even though it is very important…my son is studying medical-medicine, but I guess I just go through the syllabus, but nothing is there.”* #8, Environmentalist

All the environmentalists professed to never having specifically studied health impacts of climate change during the course of their education. On the other hand, the health professionals expressed incongruent views on climate change-health education. While one said:

*“Climate sensitive diseases, this is a part of the curriculum, we are seeing lot of diseases. And the epidemiology portion they will tell this is exacerbated during this season, this is exacerbated during this climate. That will be there in our curriculum.”* #12, Practicing physician

Another said,

*“No. So diseases we have been trained, but we have not been taught that this is so much related to the climate change. Once we completed our MBBS or MD, or during your MD we may have read it, but as a separate training or as a separate chapter, ‘these are the climate sensitive diseases’ or separate training for a week or a module, that is not part of the core curriculum.”* #3, Medical doctor/academic

Continuing education courses specific for health impacts of climate change were suggested by a few participants as potential options to bridge the gap between the environment and health. Two participants also suggested including short courses on this topic for all people working on topics related to climate change, health, adaptation and resilience.

#### Weakness in inter-sectoral information dissemination

Many of the participants mentioned having little to no awareness on climate change-health related research unless actively searching for it, pointing to the scope for improving related education and science dissemination, especially among the scientific community. Some participants were also of the opinion that awareness on health impacts of climate change in general was lacking, especially among medical professionals. Environmental risk factors were not commonly associated with health inherently, partially attributed the low scientific exposure on the topic.

*“And among doctors itself, there is not much awareness actually. How it’s going to affect, because now people are only focusing on environment change and other things actually. How we decide an impact on health, we use a lot of data. (Data) has to be there, it needs to be published.”* #6, Practicing physician

CVDs were seen as a ‘silent’ disease, with many people are not trained to look for symptoms, much less correlate them to weather conditions, all suggesting the need for improved CVD literacy and awareness on the topic. On the other hand, many participants were open to changing theirs current schools of thought on risk factors for health to include climate change, conditional to being informed by global research on the topic.

*“If the research or it’s already proven in other countries, ‘so this is a risk factor it is a good idea to add’ but [before adding anything], I think some data or there should be some routine surveillance or monitoring system should be there. … even within the medical circle, people may not be aware like how much is the contribution of this, the climate change to the heart disease or for any disease for that case… I don’t think our administrators or even our clinicians are that much thinking about the impact of climate change, and this one.”* #3, Medical doctor/academic

#### Scepticism and low awareness on non-conventional health impacts of climate change

As alluded to previously, health impacts of climate change are often not explicit, making it a challenge to research or focus the research agenda on for several reasons. One participant described how the slow pace of climate impacts leads people to think it will not immediately affect health, unless the impacts are drastic.

*“…The problem has to become so severe, like you have air pollution in Delhi, then people will act. So this climate change affects the life slowly it’s not a drastic…that is one of the reasons I feel. And slowly if you get some data and keep on generating awareness not only among the public, but also within the scientific community, then slowly things will be better.”*#3, Medical doctor/academic

For researchers, an additional challenge of convincing funders or collaborators on the health impacts of climate change also emerged. One participant described the difficulty researchers had separating environmental risks from other common health risk factors. Scepticism when attempting to research health impacts of climate change was also encountered. Confounding from other risk factors and potential ecological bias was seen as the roots of this uncertainty.

*“Maybe for six, seven years, I have been trying to do some work on climate change and environmental health. Every time I write a proposal I’m criticized largely telling that “how is it going to work?*… *And one other problem I see with the research with climate change or any environmental thing, it’s ecological effects. So people ask “how can you attribute this to only this, why not to this?”, “ Why not to lifestyle, why only to climate change?” So this direct relationship is not there.”* #4, Medical doctor/ academic

Diseases such as malaria, with historical links to stagnant water as breeding grounds, have been etched into public knowledge and further perpetrated through mass awareness campaigns, intervention programs and research. The slow developing nature of CVDs and the prevalent categorization of CVDs as solely lifestyle diseases was mentioned by many participants as potential barriers to research. The multifactorial nature of CVDs was thought to add to the difficulty of identifying climate attributable impacts, with most of the focus being on evident lifestyle contributions.

*“Non-communicable diseases, because we are not quantifying that and because of the long latent period of the incident, you’re not able to quantify directly to environment or climate change. So definitely, hypertension, cardiovascular disease, all these probably diabetes also because of the changing food pattern, but I don’t think -you cannot separate climate change from any of the health effects or any of the non-communicable diseases. Also related to stress caused by climate change.”* #2, Medical doctor/ academic

Another participant described how clinicians especially do not see the need to focus on environmental risk factors for CVDs, believing it ineffective in reducing the overall burden. One participant described how CVDs are commonly reduced to lifestyle diseases with the onus of risk management on the individual rather than a “willingness to see the invisible factors”. A few participants expressed belief and hope that the temperature-CVD association was an upcoming topic of interest for the government and public both.

*“I’m just giving my opinion. See, this heart attack and cardiovascular diseases, whatever is a well-known thing, even a common person knows that, but this climate change aspect of it, even the officials or the administrators wouldn’t know because-so, there comes the gap…So, maybe in another five years, people would really relate these two.”* #9, Environmentalist

The need for regional studies was also stressed upon as there seemed likely to be a disconnect in comparing national or global level problems with health impacts of climate change on a local level. Participants described the attitude of *“this does not affect us”* among the public when it came to climate change especially.

### Technical barriers to research

#### Insufficient resources and workforce dedicated to research

Resource allocation, especially financial, for climate change-health research was described as a barrier. Along with inconsistent funding from the government, one of the problems mentioned was lack of adequate trained personnel. This was partially linked to the need to relieve the research expectations from already over-burdened doctors. There was also a need to have trained personnel for digitalization and categorization of health data in order to create a digital state-level health database.

*“The UT of Puducherry, we do not have much data. So we need to focus and we need manpower. We do not have the funding also it’s a problem now. Sometimes they provide funds; sometimes they do not give that adequate funds.”* #1, Practicing physician/policy advisor

Some participants, referred to the low percentage of the annual budget of India allocated to health along with the need to increase this. One participant described funds earmarked for climate change-health research institutionally, along with optimism that this would lead to future research opportunities.

*“Yes, for recent years even ICMR (Indian Council of Medical Research) has called for proposals on this environment related, uh, this one. ICMR is one of the largest body which is for the research organization as well as for the academic institutes like us. So, clearly, they are given a separate block of funding for climate change and this one. That means the funds are available*.”#3, Medical doctor/academic

However, this was countered by the notion that most of the funds are directed to Central government institutes as opposed to smaller research institutes. A participant also alluded to misappropriation of research funding at an institutional level. Another participant spoke about the need to involve university students in research along with concern that most students do not get access to funding or research opportunities. There was a feeling that most students remain unaware of opportunities for funding or that funds do not ultimately reach the students aiming to conduct research. Another participant also described the prioritization of more immediate health burdens and curative research as opposed to preventative research for the directing of funds or resources. This was supported by the opinion expressed by an environmentalist on climate change being viewed as a problem for the future as opposed to the present.

*“So though we focus on vaccination and other things, but still, the budget still flows more for the curative aspects rather than the preventive part. So for instance, the climate change is more of like, you prevent this-the future heart attacks or some other diseases. You have to focus on the prevention.”* #3, Medical doctor/academic

#### Underdeveloped transdisciplinary research capacity

Alongside education, the need to build more technical capacity among researchers was also mentioned as one of the biggest challenges by participants. Despite a potential interest and willingness from researchers, the lack of training and expertise in climate change-health research was strongly expressed. This was tied in with the expressed desire for mentorship, both to facilitate increased awareness among the scientific and medical community as well as increased regional research on health impacts of climate change.

*“Yeah, more than research, I would tell it as people are aware and willing to do it, but here is more of capacity building…Let’s say if I want to work on vector borne disease, I know who to approach…but when it comes to climate change, that linking is absent. …So actually, even if I’m interested and I want to work on it, there are a lot of hurdles which has to be crossed…So I have to be given an opportunity to work on it, or I feel somebody has to mentor me to work on it. So what we call as, starting trouble, you know is there. Once I think somebody starts, we will be going into it “* #4, Medical doctor/ academic

Some participants had the belief that larger research institutes or relevant ministries could be drafted to provide training to the smaller educational institutes or local government bodies. There was a sense of “duty” attached to studying all aspects of climate change impacts for the environmentalists in Puducherry tied in with a search for a starting point.

#### Research slowed by unavailability and limited access to quality data

Participants described critical gaps in monitoring, surveillance and database development, all of which were perceived to hamper research conduction, especially for health data. First, merging health data from the many healthcare facilities within Puducherry was seen as a challenge. There was an expressed need to bring together health data for the entire UT in a single system, including public and private healthcare facilities.

*“Puducherry as so many medical colleges, but all of them do not supply data to the government. They may have their own data. That’s the problem: actually we have to integrate everyone”* #1, Practicing physician/policy advisor

Second, some participants mentioned the state-level government health-monitoring database. However, participants described this as being limited to selected diseases from all the government run primary healthcare centres, with limited information on the private sector or secondary and tertiary care hospitals. A few participants described the lack of disease-specific categorization of health outcomes, making it an added challenge in conducting health related research.

*“Cases are reported but not categorized into these sectors And as you said, this co-morbid or cardiovascular, those kind of things we do not have any data. They have a very comprehensive data. They put all the data together, (but) they don’t categorize it. Maybe now if we are very much interested in those data, we can go, collect all their data and categories for our use…They don’t have a ready data.”* #9, Environmentalist

Third, participants also perceived private medical colleges and healthcare facilities as reluctant to share data with the government, with a felt need to enhance governmental efforts to work on the state wide database. Fourth, on a related note, concerns about data quality were mentioned by several participants. Part of the reason for an unwillingness to share data by healthcare facilities was thought to be due to potentially inaccurate or poor quality data.

*“They’re all afraid of like somebody will find a fault with that. So because they don’t have manpower to look at the accurate or clean the data, okay, so somebody shares and later they find their mistake, and they will be answerable to the higher authority. So that’s the usual reason we do not to share the data, the insecurity.”* #3, Medical doctor/academic Another challenge shared was the slow, ongoing effort to digitalize the data. Participants described as feeling unmotivated to start research at the cost of manually sorting through thousands of paper records, unless there was a way to guarantee research output. This was also relate to a challenge of medical professionals being overburdened with work.

*“There is not even a digitalization…Many hospital doesn’t have digitalized MRD [medical records department]. For example, I was doing a study, retrospective study, collecting infective endocarditis data for past 10 years, there are more than 1000 files. How can I go through the 1000 files? It’s not possible.”* #13, Practicing physician

Surveillance of diseases was mentioned as ongoing work. Diabetes, hypertension, cervical cancer and other ‘notifiable’ diseases like infectious diseases were described as being under surveillance.

## DISCUSSION

This research examined mainly barriers faced in conducting climate change and health research. While we focused on Puducherry, the findings relatively remain relevant for India and can be extrapolated to other LMIC settings [33].

In recent years, there have been a lot of strides taken in the Indian policy space with pertaining to climate change and health, such as the addition of the Health Pillar to the NAPCC and the subsequent development of the NAPCCHH and mandates for the development of the State level action plans for climate change and health [17, 18]. Although the Health Pillar is a relatively recent addition (2015), we still found a substantial lack of awareness on the NAPCC as well as the health mission in general, which we present as a key area for strengthening. Knowledge of such policies, especially if they can provide a framework to support related research, is a useful tool to advance the research agenda on climate change and health [34, 35]. Health system vulnerabilities are already being seen in Puducherry and active knowledge of such policies can also be utilized by relevant stakeholders to develop resilience focused interventions. This includes communicating the severity of the problem to the policy makers, who generally lack the political will to divert resources to non-apparent problems, alluded to by the participants in this study [36, 37].

Most political efforts are thought to be focused on mitigation measures such as air pollution control, with little importance given to health adaptation and healthcare resilience. The participants believed that the health impacts of climate change were not a political priority or seen as urgent. Similar findings have been elucidated in other studies which also found public health leadership on climate change to be fragmented [25, 38]. Further efforts to inform the decision makers on the importance of health adaptation might contribute to more evidence informed climate change and health policies [39, 40]. As an added justification for health co-benefits of mitigation can be introduced through multiple pathways, including air pollution, lifestyle modification, health surveillance or research programs in development or related policies [41, 42].

We found almost unanimous support for a separate inter-sectoral body focused specifically on climate change and health. Methodological challenges in the light of limited technical knowledge and adequate inter-sectorial coordination and support for transdisciplinary capacity that we found have also been reported elsewhere [43, 44]. A recent study on the knowledge, attitudes and practices related to climate change and health among the Indian health workforce found intermediate or delayed health impacts of climate change less commonly identified [26]. This could also support the development of regional, national or even international research networks facilitating knowledge sharing and transfer, including research methodology support [44].

The compartmentalization of work within institutes or sectors was seen as a research barrier, partially due to the unclear division of responsibilities and fragmented institutional focus, as also seen in other studies [38, 44, 45]. A study examining the challenges for the Californian public health sector in climate change found the compartmentalization and lack of inter-sectorial coordination to limit work on inter-sectoral issues such as climate change and health [25]. These findings point to the need to have regular national level conferences or improved science dissemination systems to communicate climate adaptation related research or plans between and across sectors.

Our respondents had varied views regarding education on climate change or its health impacts; however, the need to improve this was clearly described. The need for strengthening capacity and education has been a common finding in several other global studies. Globally, there is a critical gap and scope for improvement in the education on health impacts of climate change, especially for medical practitioners [25, 46-54]. A study comparing medical curriculums across the world found inconsistencies between environmental changes, health and community needs, with Indian and Chinese students especially having a gap in the inclusion of planetary health in medical schools [50]. The inclusion of planetary health from an early stage for medical students leads to a more active role of physicians in educating their patients about climate risks [50, 55]. However, there is a need to validate the results in future studies given the inconsistencies in the views we found on climate change-health education. The emphasis on cure rather that prevention, which has also been shown to reduce long term healthcare costs, implying a need for Puducherry to focus on the preventative aspects, largely through education and awareness [24]. As also found in our study, these health impacts are viewed as ‘invisible’ compared to more conventional or immediate impacts, such as air pollution or extreme events [56]. This is also a commonly identified challenge to climate change and health research, accompanied by insufficient education about climate systems during the course of school or university education [26, 56].

Data barriers remain common challenges in public health research, despite efforts to facilitate improvements [57, 58]. As Puducherry has the advantage of a relatively small size and well-connected healthcare facilities, efforts need to be taken to improve a central, disease specific data collection system, incorporating all the healthcare facilities in the State [59]. Facilitating training to build local data analysis expertise would contribute to more region specific research on the topic [60]. As was also made apparent during the interviews, health impacts of climate change are a relatively new concept and not inherently associated with climate, potentially explaining the uncertainties and scepticism expressed, especially for diseases that do not warrant a visit to the doctor [61, 62]. On the positive side, the expressed desire of participants to learn more about it and make changes to the healthcare system and policies based on robust, conclusive evidence implies a willingness to adapt and implement changes in how the region tackles health impacts of climate change [49, 63]. Resource and funding constraints are one of the most common barriers to public health research, especially in LMICs and there remains a critical need to address this gap [64].

At present, little is known on CVD impacts of climate change in India. Our related study from Puducherry found a high attributable burden of non-optimal temperature to CVD mortality, suggesting a need for similar studies from around the country [65, 66]. The CVD specific challenges we identified here are comparable to the general health challenges. Awareness among the medical community on the environmental risk factors of CVDs will be instrumental in furthering this research agenda, while awareness among policy makers will help raise the political prioritization of CVD impacts of climate change [24, 67, 68].

## Limitations

This study had a few limitations. First, the sample was restricted to Puducherry district and not representative of the entire Union Territory of Puducherry, much less India as a whole, although the projected population for Puducherry is 1.25 million in 2021, comparable to a few smaller countries or global regions [69]. The results might thus only reflect the studied context and participants. Secondly, while we chose to focus on the medical community and DSTE representatives working on climate change, we could not include the experiences and perspectives of other public health professionals or stakeholders. Third, we do not highlight the opportunities for increasing research on climate change and health as many of these are very often interconnected with barriers. However, we do discuss potential recommendations given by stakeholders. Finally, our sample size was also limited by the ongoing COVID-19 measures. Nonetheless, the results of this study could be useful for the research community and policy makers alike to strengthen climate change and health research and engagement.

## Conclusion

There is a great need to fill the gap in research on the impacts of climate change on various health outcomes in India, especially in light of the vulnerabilities it faces. By highlighting some crucial barriers to environmental health research faced by relevant professionals, we present potential intervention points for consideration. Insufficient awareness on health impacts of climate change and perceived need to improve research capacity through collaborative work; and challenges in data availability emerged as the largest barriers to conducting research on this topic in Puducherry. We outlined the gaps and scope for addressing these through improved policy awareness; informed leadership and evidence informed climate change and health policies; research capacity strengthening and transdisciplinary research and communication network; improved education on climate change and health on all levels; and addressing data barriers in climate change through improved monitoring and evaluation systems. The key findings could contribute to supporting and strengthening evidence-informed climate resilient healthcare systems. In addition, it would also serve to inform and strengthen the research and institutional support for environmental health research in the future both in India and globally.

## Data Availability

All relevant data from this study has been included in the Supplementary material. As this is a qualitative study with a small number of key informants, making the full dataset and interview transcripts available to a wider audience could potentially breach the confidentiality commitment made to the participants during the process of obtaining informed consent as well as to the ethics committees that approved this study. Therefore, this data will not be made available.

## DECLARATIONS

### Author contributions

S.S, M.R, M.A.D, J.U and G.C conceptualized and planned the study. S.S and R.L acquired and provided access to the data. R.L facilitated the interviews. S.M and S.S designed the study. T.L validated the codes and codebook. O.C, S.S and S.M conceptualized and structured the framework. S.S wrote the main manuscript with inputs from all authors. The final manuscript has been revised by all authors.

### Informed consent

This study was approved by the Institute Ethics Committee (Human Studies) of the Indira Gandhi Medical College and Research Institute (A Government of Puducherry Institution); No. 318/IEC-31/IGM&RI/PP/2021 and by the Ethics Committee Northwest and Central Switzerland (EKNZ); Statement ID-AO_2020_00034. The methodology used in this project abided by the principles laid out in the Declaration of Helsinki and the COREQ checklist.

All participants were verbally explained the project and its objectives as well as being provided information sheets. All participants were made aware of their right to refuse participation at any point prior to publication of the study. Signed informed consent was obtained from all participants prior to the interviews, with participants retaining one copy.

## Acknowledgements

The authors would like to extend their sincere gratitude to all the participants who made this study possible.

